# An electronic cigarette pod system delivering 6-methyl nicotine, a synthetic nicotine analog, marketed in the United States as “PMTA exempt”

**DOI:** 10.1101/2023.11.21.23298778

**Authors:** Sven E. Jordt, Sairam V. Jabba, Peter Silinski, Micah L. Berman

## Abstract

As of April 14, 2022, the United States Food and Drug Administration (FDA) has been authorized to regulate tobacco products containing nicotine from any source, including synthetic, requiring manufacturers to submit a premarket tobacco product application (PMTA). A recent report by the World Health Organization (WHO) warned that non-nicotine tobacco alkaloids or other synthetic nicotine analogs could be used by manufacturers to bypass regulatory schemes focusing on nicotine alone. From October 2023 on, vape stores in the United States started selling a new electronic cigarette pod system, named Spree Bar, advertised as “PMTA exempt”, with youth-appealing flavors and advertising. The products are marketed as containing “Metatine”, a trademarked name for 6-methyl nicotine, a synthetic nicotine analog patented by a Chinese electronic cigarette manufacturer. Here we used liquid chromatography-mass spectrometry (LCMS) to confirm the presence of a chemical species with the molecular weight of 6-methyl nicotine in Spree Bar e-liquids. The FDA needs to determine whether, in its view, 6-methyl nicotine is a form of “nicotine” within the meaning of the Tobacco Control Act, or whether 6-methyl nicotine can be regulated as a drug under the Federal Food, Drug, and Cosmetic Act (FDCA).

## Background

In February 2021, the popular vaping company, PuffBar, started selling disposable e-cigarettes in the United States containing synthetic nicotine, claiming to be exempt from federal and state laws regulating products containing tobacco-derived nicotine. ^1^ United States lawmakers responded to the emerging synthetic nicotine market by clarifying that synthetic nicotine products are subject to the U.S. Food and Drug Administration’s regulatory regime for tobacco products. ^1-3^ As of April 14, 2022, FDA has been authorized to regulate tobacco products containing nicotine from any source, including synthetic, requiring manufacturers to submit a premarket tobacco product application (PMTA). ^3^

While many countries have updated their tobacco product laws to cover synthetic nicotine, or are in the process of doing so, a recent World Health Organization (WHO) report warned that non-nicotine tobacco alkaloids or other synthetic nicotine analogs could be used by manufacturers to bypass regulatory schemes focusing on nicotine alone. ^2^

### Spree Bar, a “PMTA exempt” electronic cigarette system

As of October 2023, this is not just a hypothetical concern. Vape stores in the United States started selling a new electronic cigarette pod system, named Spree Bar, advertised as “PMTA exempt” (figure 1). ^4^ Announced by the company, Charlie’s Holdings Inc., in March 2023, Spree Bar products are promoted through convenience store advertising and web channels, using youth- and young adult-appealing artificial intelligence-generated characters (figure 1). The products are marketed as containing “Metatine”, a trademarked name for a synthetic nicotine analog (figure 2). ^5-7^ The company states that:

**Figure 1.**
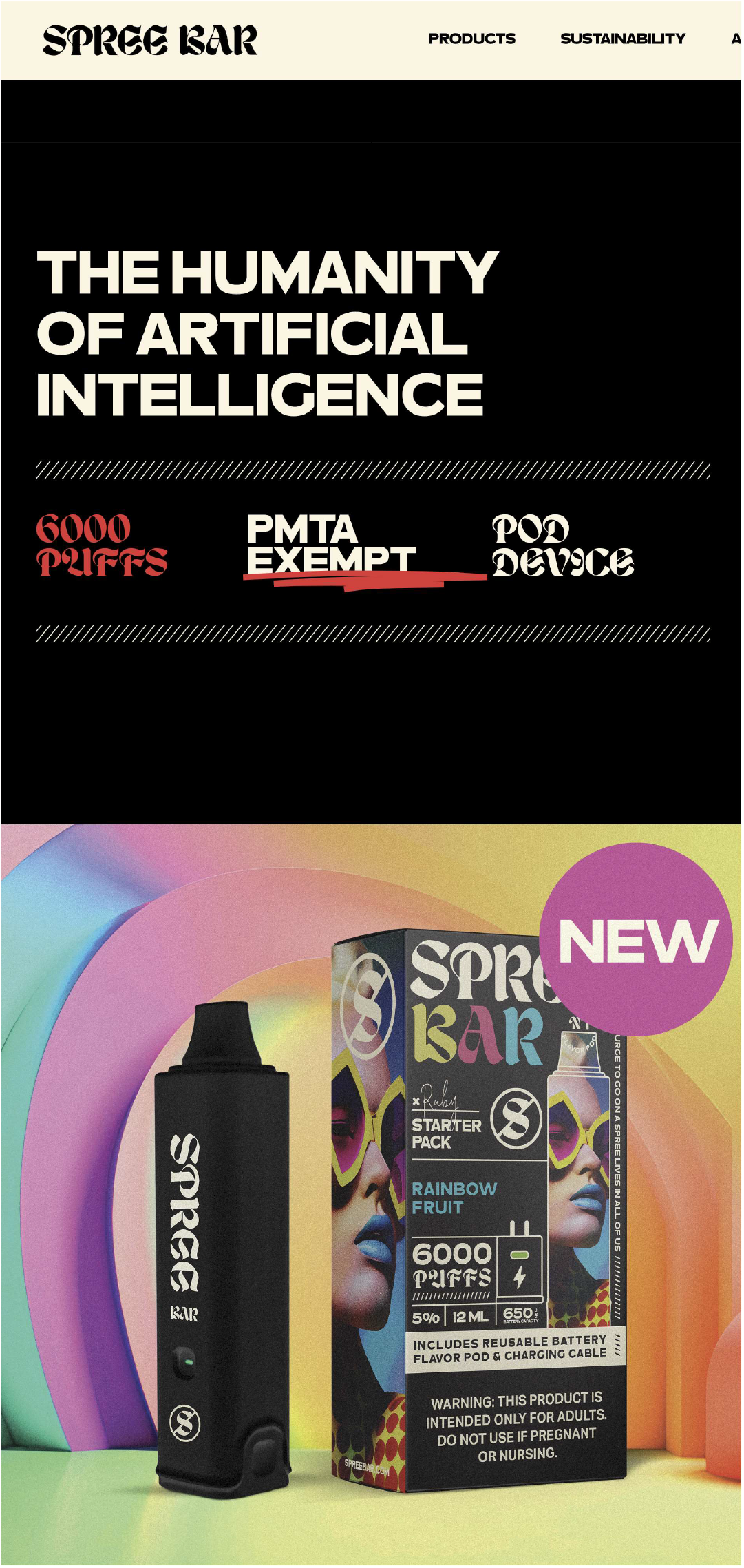
Screen shot of the Spree Bar website home page taken on 10/08/2023, with a statement claiming that the products are “PMTA exempt”. ^4^

**Figure 2.**
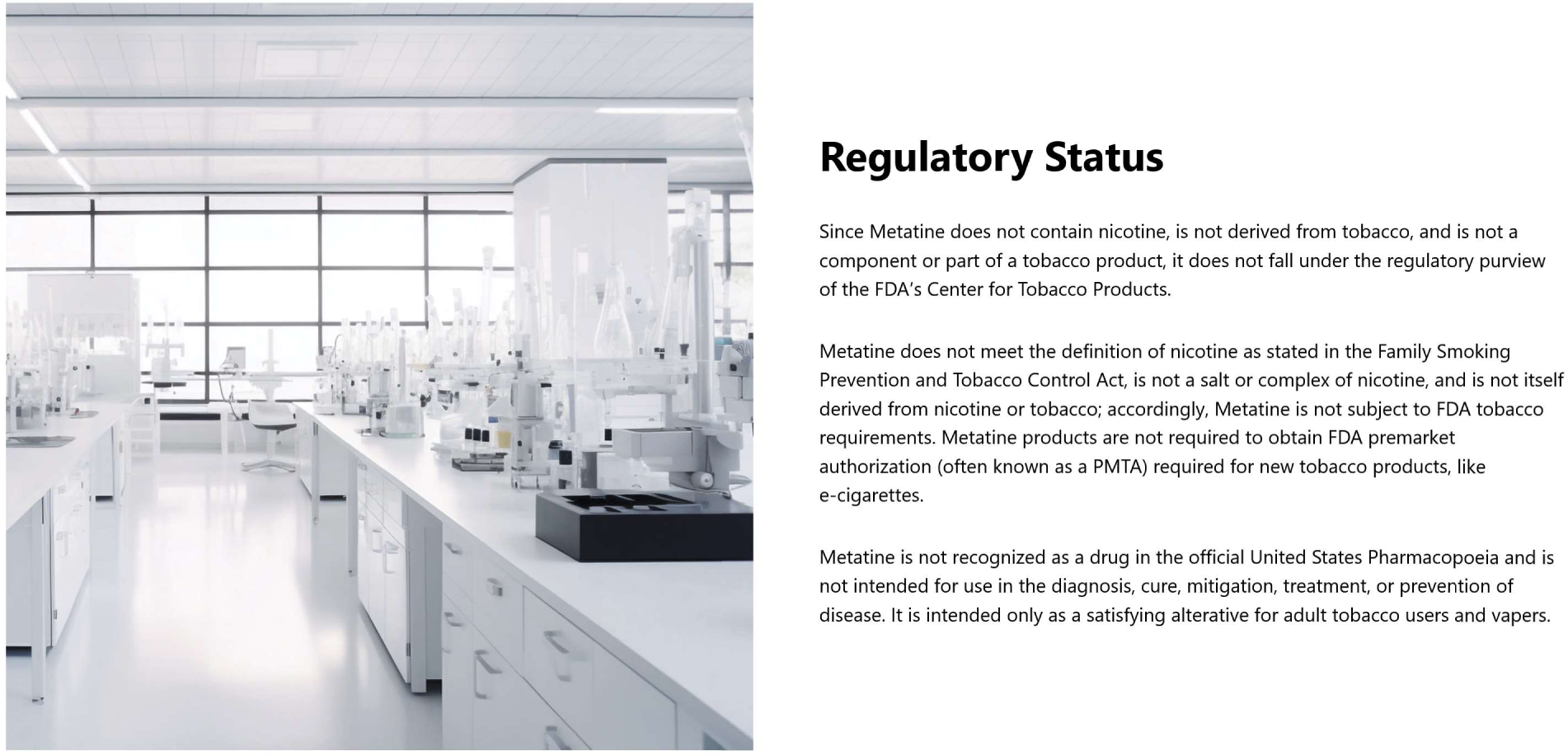
Section of a screen shot of the Metatine website home page taken on 11/21/2023, stating that products containing the active ingredient, 6-methyl nicotine, do not meet the definition of “tobacco product” in the Federal Food, Drug and Cosmetic Act, and do not require a PMTA to be sold in the US. ^8^

“***SPREE BAR is pre-filled with flavored e-liquid containing proprietary Metatine™***. *Metatine is a synthetically derived molecule that is structurally similar to, but chemically different from, other vaping alkaloids. Although Metanine produces the same sensation as nicotine and may also be addictive, Metatine is not made or derived from tobacco or nicotine, and Metatine does not consist of or contain nicotine from any source*.

*SPREE BAR products containing Metatine (and no tobacco-derived materials or nicotine) do not meet the definition of a “tobacco product” in the Federal Food, Drug, and Cosmetic Act and do not require a Pre-Market Tobacco Application (PMTA) to be sold in the US*.”. ^4^

### “Metatine” (6-Methyl nicotine) as a nicotine replacement

On a website dedicated to “Metatine” the company states that:

“*The substance Metatine™ has the International Union of Pure and Applied Chemistry (IUPAC) designation 2-methyl-5-[(2S)-1-methylpyrrolidin-2-yl]pyridine …*.”.^8^

2-methyl-5-[(2*S*)-1-methylpyrrolidin-2-yl]pyridine is synonymous with (*S*)-6-methyl nicotine (Chemical Abstracts Number: CAS 13270-56-9) (figure 3A). ^9^ 6-methyl nicotine is a structural analogue of nicotine, with a methyl group substitution at position 6 of nicotine’s pyridine ring (figure 3A). Recently awarded patents describe new synthesis protocols to generate 6-methyl nicotine from petrochemical precursors. ^10-12^ One such patent was assigned to Shenhzen Zinwi Biotech, a leading e-liquid company that also sponsored a toxicological study at Guangdong Pharmaceutical University. This study reported that 6-methyl nicotine has increased cytotoxicity compared to nicotine in a permanent human bronchial epithelial cell line (BEAS-2B), but milder effects on the upregulation of lung cancer-related proteins. ^10 13^ The authors conclude “*that new electronic cigarettes with 6-MN might offer some advantages over conventional electronic cigarettes containing nicotine*”, providing evidence that the compound, 6-methyl nicotine (abbreviated as 6-MN) is considered for addition to e-cigarettes. ^13^

**Figure 3.**
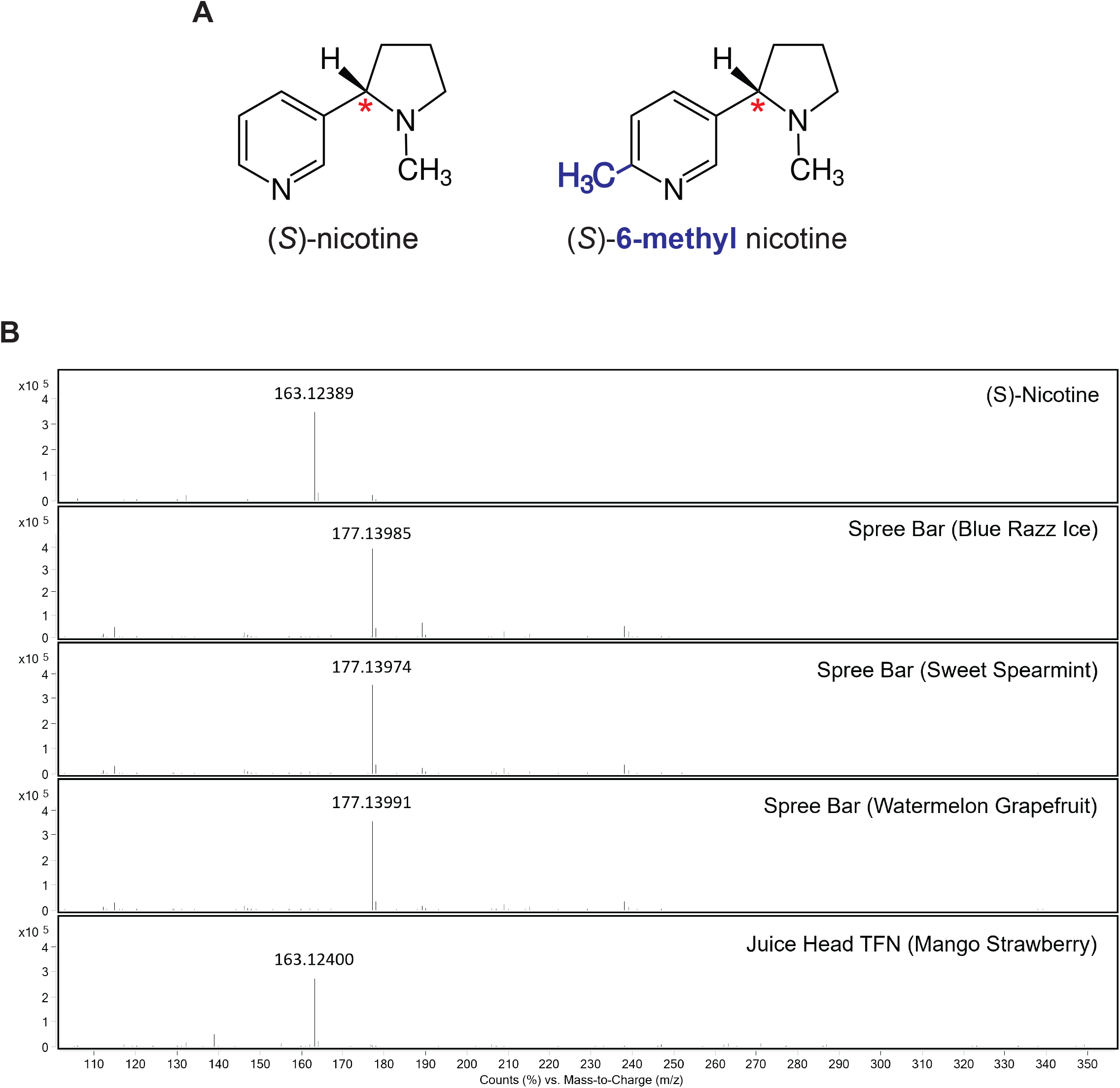
**A**. Chemical structures of (*S*)-nicotine (left), the major form of nicotine in tobacco, and (*S*)-6-methyl nicotine (right), trademarked as “Metatine”, with a methyl group substitution (blue) at position 6 of the pyridine ring. The red asterisk indicates the chiral center of nicotine. **B**. Mass spectra from extracted ion chromatograms for pure (*S*)-nicotine and four e-liquids. The major observed ion in each spectrum corresponds to the expected [M+H]^+^ ion for either nicotine (163.1230 m/z) or 6-methylnicotine (177.1386 m/z).

Another patent, assigned to Shanghai Lingnuo Biotech Co Ltd includes human studies in which e-cigarette users were asked to rate the “throat hit” and other psychophysical qualities of e-vapor containing 6-methyl nicotine. ^11^ 1 mg/ml 6-methyl nicotine was found to be as satisfying and producing a similar “throat hit” as 3 mg/ml nicotine, concluding that “6-methyl nicotine can be used for replacing nicotine in electronic aerosolization”. ^11^

Charlie’s Holdings Inc., the vendor of Spree Bar also claims that: “*Metatine products are exempt from nicotine excise taxes in many states, which can result in massive savings for retailers and consumers. Metatine can be combined with many flavors and is legal to sell in nearly all 50 states*.”. ^14^

Spree Bar products and Metatine are discussed in Reddit posts, including initial user reports, suggesting increasing awareness by e-cigarette users in the United States. ^15 16^ The authors of the present study purchased Spree Bar products on 10/07/2023, receiving delivery on 10/17/2023, confirming availability in the United States. ^17^ An abstract presented by a US-based chemical analytical services company at a recent tobacco industry conference describes a method for the quantification of 6-methyl nicotine and detected its presence in e-liquids, confirming efforts by the industry to market such products. ^18^

### Chemical analysis

Chemical analysis of three Spree Bar e-liquids (Sweet Spearmint, Blue Razz Ice, Mango Grapefruit), a pure (*S)*-nicotine standard and a commercial e-liquid known to contain synthetic nicotine (Juice Head TFN Mango Strawberry) was carried out using high-performance liquid chromatography mass spectrometry (LCMS). The presence of either (s)-nicotine or 6-methylnicotine was investigated using extracted ion chromatograms (EIC) for the theoretical [M+H]^+^ ions for the target compounds (163.1230 m/z for nicotine, and 177.1386 for 6-methylnicotine). The EIC peaks corresponding to these ions were found to co-elute near the beginning of the gradient, consistent with the compounds being relatively polar. Mass spectra for the resulting EIC peaks for the standard and e-liquids are shown in figure 3B. In the pure (*S)*-nicotine control and the commercial e-liquid (Juice Head) declared to contain synthetic nicotine, a signal near 163 m/z was observed, consistent with the presence of nicotine. No signal near 177 m/z was observed in the standard or Juice head sample, suggesting the absence of 6-methylnicotine. Conversely, in the Spree Bar liquids, a signal near 177 m/z was observed, consistent with the presence of 6-methylnicotine. No signal near 166 m/z was observed in the Spree liquid samples, suggesting the absence of nicotine from these samples. The mass accuracy of the observed ions were all within 7 ppm of the theoretical values.

### 6-Methyl nicotine: Regulatory challenges and solutions

Are products containing 6-methyl nicotine, differing from nicotine by a methyl group, subject to the PMTA requirement and the other provisions of the U.S. Tobacco Control Act (TCA), or are they indeed “PMTA exempt”? The TCA, as amended in 2022, now covers products “made or derived from tobacco, or containing nicotine from any source.”. ^19^ The TCA does not further define the term “nicotine” in scientific terms, so it will be up to the FDA to determine whether, in its view, 6-methyl nicotine is a form of “nicotine” within the meaning of the TCA. If the FDA concludes that it is, and it takes enforcement action against Spree Bar or other similar products, the manufacturers may challenge that determination in court.

However, should the FDA (or a future court decision) conclude that 6-methyl nicotine is not “nicotine” within the meaning of the TCA, that does not mean that products containing 6-methyl nicotine are necessarily exempt from all FDA regulation. The proclamations on Spree Bar’s website that Metanine “produces the same sensation as nicotine” and “may also be addictive,” combined with the similarity in chemical form to nicotine and evidence of similar physiological effects, strongly suggest that the FDA could alternatively seek to regulate the 6-methyl nicotine in Spree Bar as a drug under the Federal Food, Drug, and Cosmetic Act (FDCA). ^20^ If the FDA determines that 6-methyl nicotine is being marketed as a drug intended to produce particular physiological effects (the FDCA defines drugs in part as “articles (other than food) intended to affect the structure or any function of the body”) ^21^, then it cannot be legally sold without the manufacturer first obtaining FDA approval.

When companies (inaccurately, in our view) market products like Spree Bar as exempt from FDA regulation, it reduces confidence in the FDA’s regulatory scheme and undermines the FDA’s efforts to protect public health. The FDA should quickly announce its policy with respect to such products and take appropriate enforcement action. Congress might also consider clarifying that products containing non-nicotine tobacco alkaloids or synthetic nicotine analogues are tobacco products under U.S. law, by amending the TCA to expressly say as much.

## Methods

### High-Performance Liquid Chromatography Mass Spectrometry (LCMS)

The reference sample for pure S-nicotine (Sigma-Aldrich, St. Louis, MO, USA) was diluted 10,000X into water, and electronic cigarette liquids (Spree Bar, Juice Head TFN) were diluted 1000X into water prior to analysis. The samples were then analyzed on an Agilent 6224 TOF LC-MS system consisting of a 1200 HPLC (degasser, binary pump, thermostated column compartment, diode array detector (DAD)) coupled to a 6224 accurate-mass time-of-flight mass spectrometer. The mass spectrometer was equipped with a Dual ESI source for accurate mass determinations. Positive-ion mass spectral data were acquired in full-scan mode over the range of 60-1700 m/z using the following source parameters: gas temperature 325 °C, gas flow 11 L/min, nebulizer pressure 33 psig, VCap 3500 V, and fragmentor voltage 150 V. HPLC separations were achieved on a Phenomenex Kinetix C18 column (3 × 30 mm, 2.6 μ) using a linear gradient of mobile phase B (acetonitrile with 0.1% formic acid) in MP A (0.1% formic acid), a flow rate of 0.5 mL/min, and a column temperature of 40 °C.

## Data Availability

All data produced in the present study are available upon reasonable request to the authors

## Disclaimer

The funding organization had no role in the design and conduct of the study; the collection, management, analysis, and interpretation of the data; the preparation, review, or approval of the manuscript; nor in the decision to submit the manuscript for publication. The content is solely the responsibility of the authors and does not necessarily represent the views of National institutes of Health or the Food and Drug Administration.

## Author Contributions

SEJ and MLB conceptualized and designed the study and wrote the first draft of the paper; SVJ provided advice on product choice and figures design. PS carried out the chemical analysis of the e-cigarette products. SEJ, SVJ, PS and MLB contributed to revision of the manuscript. All authors critically reviewed, edited, and approved the final draft before submission. SEJ attests that all listed authors meet authorship criteria and that no others meeting the criteria have been omitted.

